# Evidence of accelerated epigenetic aging in patients diagnosed with coronary artery disease: Results of the LipidCardio study

**DOI:** 10.1101/2020.02.23.20026906

**Authors:** Verena Laura Banszerus, Valentin Max Vetter, Maximilian König, Ulf Landmesser, Ilja Demuth

**Affiliations:** Lipid Clinic at the Interdisciplinary Metabolism Center, Charité - Universitätsmedizin Berlin, corporate member of Freie Universität Berlin, Humboldt-Universität zu Berlin and the Berlin Institute of Health, Berlin, Germany; Medical Department, Division of Nephrology and Internal Intensive Care Medicine, Charité -Universitätsmedizin Berlin, Berlin, Germany; Department of Cardiology, Campus Benjamin Franklin (CBF), Charité - Universitätsmedizin Berlin, Berlin, Germany; Berlin Institute of Health (BIH), Deutsches Zentrum für Herzkreislaufforschung (DZHK), Parter Site Berlin, Berlin, Germany; BCRT - Berlin Institute of Health Center for Regenerative Therapies, Charité - Universitätsmedizin Berlin, Berlin, Germany

**Keywords:** Epigenetic clock, coronary heart disease, coronary artery disease, ischemic heart disease, cardiovascular risk factors, DNA methylation (DNAm) age acceleration

## Abstract

DNA methylation (DNAm) age acceleration, defined as the deviation of chronological and epigenetic age determined by an epigenetic clock, has been proposed as a biomarker of biological aging. To address the above hypothesis in the context of cardiovascular disease, we evaluated whether patients (N=827, mean chronological age: 69.82±11.01 years, DNAm age: 71.91±16.11, residual DNAm age acceleration: 0.00±9.65 years), who were diagnosed with obstructive coronary artery disease (CAD) by coronary angiography were aged prematurely, i.e. had an increase in the DNAm age acceleration, in comparison with patients for whom obstructive CAD was ruled out (controls).

Stratified analysis yielded a significant acceleration in DNAm age (determined by a seven cytosine-phosphate-guanine epigenetic clock) in patients diagnosed with obstructive CAD, defined by at least one >50% coronary stenosis (N=588, rDNA age acceleration=0.58±9.47, corrected p= 2.05⨯10^−3^) compared to control subjects (N=145, residual (r)DNAm age acceleration= -3.11±10.51 years). Moreover, rDNAm age acceleration was significantly associated with systolic blood pressure (ß=0.069, 95% CI 0.027 – 0.112, p= 1.44⨯10^−3^), sex (ß=-2.438, 95% CI -4.591 - -0.285, p= 2.65⨯10^−2^), estimated glomerular filtration rate (eGFR, ß=0.040, 95% CI 0.011 – 0.069, p= 6.87⨯10^−9^) and smoking status (ß=-8.538, 95% CI -10.772 - -6.303, p= 2,45⨯10^−13^).

Across studies, assessing CAD and its risk factors in the context of epigenetic age acceleration findings are remarkably inconclusive. While the here employed seven-cytosine-phosphate-guanine epigenetic clock suggests premature biological aging in CAD patients, compared to controls without coronary stenosis, its association with cardiovascular risk factors was limited.

## Introduction

Chronological age is strongly associated with functional and cognitive capacity, morbidity and mortality and is the most informative cardiovascular risk factor. However, chronological age is unable to depict the remarkable heterogeneity of biological aging rates observed in peers. Moreover, increasing life expectancy and the demographic shift towards a chronological older world population (UN Department of Economics and Social Affairs, 2019), foster the demand for clinically meaningful biomarkers of biological aging progressively.

Due to its accuracy to predict chronological age, the epigenetic clock has been suggested as an alternative biomarker of aging in 2013 (Hannum, Guinney, Zhao, Zhang, & Hughes, 2013; Horvath, 2013). The epigenetic clock parameter, DNA methylation (DNAm) age is calculated by the weighted DNAm fraction of a number of cysteine-phosphate-guanine (CpG) nucleotides, selected by penalised regression analysis (Hannum et al., 2013; Horvath, 2013). We have previously shown that DNAm age (measured by a seven-CpG epigenetic clock trained in the LipidCardio cohort) is correlated with chronological age independent of the established biomarker of aging, relative telomere length, in the LipidCardio cohort (Banszerus, Vetter, Salewsky, König, & Demuth, 2019). Whether the discrepancy of DNAm age (determined by a seven-CpG epigenetic clock trained in a mostly health cohort of the Berlin Aging study (Vetter et al., 2018)) and chronological age, termed *DNAm age acceleration*, is able to distinguish decelerated, age-appropriate and accelerated biological ageing, across different clinical aging-phenotypes, including cardiovascular disease (CVD) and coronary artery disease (CAD) remains to be validated further and will be examined in this cross-sectional analysis of the LipidCardio cohort.

Coronary artery disease (coronary/ischemic heart disease) is a chronic cardiovascular disease, which is characterised by the progressive atherosclerotic plaque formation reducing the diameter of coronary arteries, eventually leading to chronic ischemia of the myocardium, acute coronary syndrome, heart failure or cardiovascular death. While improvements in health behaviour (including balanced diet, physical activity, reduced smoking), efforts to manage cardiovascular risk factors (including hypertension, dyslipidaemia) and advances in the medical treatment of coronary syndromes have reduced the incidence of CAD and CAD mortality, CAD remains the most common cause of death in Germany and globally (Busch & Kuhnert, 2017; Robert Koch Institut, 2015; Wang et al., 2016; Wilkins, Wilson, Wickramasinghe, & Bhatnagar, 2017).

The hypothesis of an association of the epigenetic clock and CVD, CAD, and its risk factors, has been assessed by a number of recent studies, yielding inconclusive results: Perna et al. studied a subcohort of 1862 participants of the population-based ESTHER cohort (chronological age: 62.5±6.6 years at baseline, 57.2% female) (Perna et al., 2016). DNAm age acceleration was determined by the difference of the Horvath estimate of biological age and the chronological age, adjusted for batch effects, sex, chronological age and leukocyte distribution. A cox proportional hazard ratio analysis revealed a 20% increased risk (95% CI 0.98-1.43) for CVD mortality per five-years increase in DNAm age acceleration. Interestingly, no association of CVD mortality and DNAm age acceleration was reported, when the DNAm age acceleration was determined by the difference of the Hannum estimate and the chronological age.

Similarly, Lind et al. reported a clock-dependent association of the DNAm age acceleration (determined by the difference of the Horvath estimate and the chronological age) and incident CVD, adjusted for sex, body mass index (BMI), diabetes mellitus status, high density and low density lipoprotein cholesterol (HDL-C and LDL-C) levels, systolic blood pressure and smoking (HR 1.033, 95% CI 1.004-1.063, p= 2.4⨯10^−2^) (Lind, Ingelsson, Sundstr, Siegbahn, & Lampa, 2017). Moreover, a one-year increase in Horvath’s DNAm age estimate was associated with a 4% increase in risk to develop CVD among the 832 participants of the Prospective Study of the Vasculature on Uppsala Seniors (PIVUS) study (chronological age: 70.20±0.15 years, 50% female), during 10-year follow-up, while no association was detected between incident CVD and DNAm age acceleration, determined by subtracting the chronological age from the Hannum estimate (Lind et al., 2017).

Roetker et al. examined the association of DNAm age acceleration (determined by the residuals resulting from the regression of the Horvath or the Hannum epigenetic age estimate on the chronological age) and carotid intima-media thickness, as well as the incidence of cardiovascular events during a median follow-up of 21 years in 2,543 participants of the Atherosclerosis Risk in Communities (ARIC) study (mean chronological age: 57±6 years, 65% female) (Roetker, Pankow, Bressler, Morrison, & Boerwinkler, 2018). Both, Horvath (p=0.004) and Hannum (p=0.002) DNAm age acceleration were independently associated with a 0.01 mm increase in mean carotid intima-media thickness per 5-year increase in the DNAm age acceleration and risk of fatal CAD (HR 1.17, 95% CI 1.02-1.33 and HR 1.17, 95% CI 1.04-1.44, respectively), adjusted for educational attainment, clinical risk factors, lifestyle factors and BMI, as well as peripheral arterial disease, myocardial infarction (Hannum DNAm age acceleration only) and heart failure (Roetker et al., 2018). Moreover DNAm age acceleration was associated with CVD risk factors including BMI, a sport index (Hannum DNAm age acceleration only), HDL-C (Horvath DNAm age acceleration only), diabetes mellitus status and smoking (Roetker et al., 2018).

In contrast, Horvath et al. observed neither a predictive association of the extrinsic epigenetic age acceleration (EEAA, measured by the residuals of the regression of the Horvath estimate on the chronological age) nor of the intrinsic epigenetic age acceleration (IEAA, residual EEAA adjusted for CD8+ T cells, plasma B cells, CD4+ T cells, NK cells, monocytes, granulocytes) with incident CAD among the 1462 postmenopausal women of the Woman’s Health Initiative (WHI, chronological age: 63 years, ranging from 50-80 years) (Horvath, Gurven, Levine, Trumble, Kaplan, Allayee, Ritz, Chen, Lu, Rickabaugh, Jamieson, Sun, Li, Chen, Quintana-murci, et al., 2016). However, EEAA was weakly associated with CAD risk factors, like hypertension, triglyceride levels, reduced HDL-cholesterol and diabetes mellitus.

Moreover, no association was found between the IEAA measured by the Hannum (HR 1.08, 95% CI 0.97-1.21, p=0.17) or Horvath (HR 1.00, 95% CI 0.90-1.12, p=0.93) estimate and CVD mortality, in the study of 2,818 participants of the Melbourne Collaborative Cohort Study by Dugué et al. (chronological age: 59.0±7.6 years, 49% female) (P. Dugué et al., 2018).

While the above studies employed the original Horvath or the Hannum epigenetic clock, which are based on the methylation status at 353 and 71 CpGs respectively, alternative clocks have been described based on different training populations, choices of tissue and laboratory analytic approaches (Vidal-Bralo, Lopez-golan, & Gonzalez, 2016; Vidal-bralo, Lopez-golan, Gonzalez, & Gonzalez, 2017; Weidner et al., 2014).

Here we employ the epigenetic clock suggested by Vidal-Bralo et al., originally included the methylation status of eight most informative CpGs, which was adapted and further reduced to a seven-CpGs epigenetic clock by Vetter et. al., (to determine the epigenetic age of the population-based Berlin Aging Study II cohort; BASE-II, N=1,895, about 50% female, chronological age of younger subcohort: 28.8±3.1 year, N=424, chronological age of older subcohort 68.7±3.7 years, N=1,471) (Demuth et al., 2019; Vetter et al., 2018; Vidal-Bralo et al., 2016; Vidal-Bralo et al., 2017). Whether the seven-CpGs epigenetic clock is able to detect the heterogeneity of epigenetic aging rates in the context aging-associated disease, like CAD and its risk factors has not been assessed previously. To address the above hypothesis we evaluated cross-sectionally, whether patients of the LipidCardio cohort (N=1005, 30.1% female, chronological age: 72.03 ± 11.07years) with an angiography-confirmed obstructive CAD were aged prematurely (assessed by the seven-CpGs epigenetic clock), in comparison with patients with non-obstructive CAD and patients with normal angiogram.

Furthermore, we assessed the association of the cardiovascular risk factors: systolic blood pressure, HDL-C and triglyceride levels, creatinine clearance, diabetes mellitus type II status, lipoprotein a, body mass index (BMI), physical activity, alcohol and nicotine consumption with DNAm age acceleration and placed our findings in the context of previous studies, identified by an inclusive literature review.

## Material and Methods

A detailed description of the LipidCardio study and the cohort profile has been published previously (König et al., 2019). In summary, patients aged 18 years and above, who underwent diagnostic cardiac catheterization for coronary angiography at the department of cardiology at Campus Benjamin Franklin, Charité – Universitätsmedizin Berlin, between October 2016 and March 2018, were eligible for inclusion, independent of their diagnosis and after providing written informed consent. Patients with acute troponin-positive coronary syndrome were not eligible for inclusion. Aim of the observational study was to collect patients’ clinical data inclusively, enabling cross-sectional association analysis with a special focus on CVD and its risk factors. The coronary angiography was performed by a trained cardiologist according to the standard protocol and documentation routine employed at the unit and allowed to allocate patients into three groups: (1) controls with normal angiogram and without evidence of atherosclerosis (2) non-obstructive atherosclerosis and CAD with a lumen reduction below 50% and (3) clinical obstructive CAD with a lumen reduction exceeding 50%. The study was conducted in accordance with the Declaration of Helsinki and approved by the ethics committee of the Charité - Universitätsmedizin Berlin, Germany - approval number EA1/135/16 (27th June 2016).

### Determination of the residual DNAm (rDNAm) age acceleration

Blood was drawn during the cardiac catheterization for coronary angiography, either via a peripheral intravenous access or from the radial or the femoral artery sheath post-heparin administration for the purpose of routine laboratory testing, leukocyte DNA extraction and storage. Whole blood EDTA samples were frozen at -80°C following collection to allow for a one-batch leukocyte DNA isolation with the sbeadex livestock kit (LGC Genomics GmbH, Berlin Germany) in accordance with the manufacture’s protocol.

The percentage DNAm status was determined at the CpG dinucleotide positions cg09809672, cg02228185, cg19761273, cg16386080, cg17471102, cg24768561 and cg25809905 (as well as cg10917602) following a laboratory protocol by Vidal-Bralo el al. adapted as described by Banszerus et al. and Vetter et al. (Banszerus et al., 2019; Vetter et al., 2018; Vidal-Bralo et al., 2016; Vidal-bralo et al., 2017). Briefly, 500ng leukocyte DNA was bisulfite converted, employing the EZ-96 DNA Methylation-Lightning Kit (ZYMO Research, Irvine, CA, USA), according to the manufacture’s protocol. DNA was amplified by multiplex polymerase chain reaction (mPCR) and enzymatically cleaned, before undergoing multiplex single nucleotide primer extension (mSNuPE) followed by a cleaning step. Oligonucleotides for mPCR and mSNuPE were designed according to Vidal-Bralo et al. (Vidal-Bralo et al., 2016; Vidal-bralo et al., 2017). A zero control of HPLC water was run alongside the samples on each multiplex plate during the entire procedure and analysed alike the samples in a 3730 DNA Analyser (Applied Biosystems, Waltham, MA, USA). GeneMapper software package 5 (Thermo Fisher Scientific, Waltham, MA, USA) was employed to assess spectra for measurement quality and to determine peak height rations to determine the DNAm fraction at each CpG dinucleotide, as suggested by Kaminsky el al. (Kaminsky, Assadzadeh, Flanagan, & Petronis, 2005). Subsequent statistical analysis was performed using SPSS Statistics 25 by IBM (Armonk, NY, USA).

The below seven-CpGs epigenetic clock formula for epigenetic age estimation, which was trained in the predominantly healthy cohort of the BASE-II study by Vetter et al. (who adapted the choice of CpGs and a laboratory protocol by Vidal-Bralo et al.), was employed to calculate the epigenetic age in the LipidCardio patients (Vetter et al., 2018; Vidal-Bralo et al., 2016; Vidal-bralo et al., 2017):

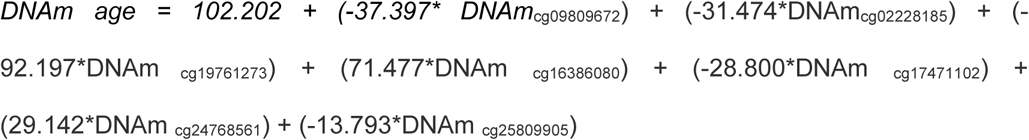

rDNAm age acceleration, was estimated by the residuals of regressing the DNAm age on the chronological age (which was determined calculated by subtracting the date of inclusion from the date of birth, divided by 365.25 days, taking leap years into account) (Rosen et al., 2018).

### Statistical analysis

Available case analysis was performed. The subcohort for which rDNAm age acceleration and CAD status was available, was stratified according to their angiographically-confirmed CAD status: (1) patients with normal angiogram, (2) non-obstructive CAD with an atherosclerotic lumen reduction below 50% and (3) obstructive clinical CAD with a lumen reduction exceeding 50% for the purpose of the descriptive statistics and the subsequent correlation. The difference of the rDNAm age acceleration across patients allocated to the three above CAD status was assessed by a student’s t-test, and a linear regression analysis adjusting for the cardiovascular risk factors including systolic blood pressure, HDL-C level, triglycerides, diabetes mellitus type II, estimated glomerular filtration rate (eGFR) determined by Cockcroft-Gault equation, lipoprotein (a) levels >107 nmol/L, BMI, Rapid Assessment for Physical Activity (RAPA) questionnaire, alcohol consumption (yes/no) and current smoking behaviour (yes/no). Further we assessed for an association of the DNAm fraction at the seven single CpGs of interest with the CAD status by linear regression analysis. A p-value of 0.05 or lower was defined as statistically significant.

### Literature Review

PubMed was searched systematically, to identify original studies, which assessed DNAm age acceleration and CAD and its risk factors, using the search term “epigenetic clock” in combination with “coronary artery disease”, “coronary heart disease”, “systolic blood pressure”, “HDL cholesterol”, “triglycerides”, “creatinine clearance”, “glomerular filtration rate”, “diabetes mellitus type II”, “lipoprotein (a)”, “body mass index”, “obesity”, “physical activity”, “alcohol” or “smoking”.

## Results

Data on rDNAm age acceleration and CAD status was available for 827 LipidCardio participants (chronological age= 69.82±11.01 years, 30.2% female). Obstructive CAD with a lumen reduction exceeding 50% was confirmed angiographically in 71.1% of the patients (chronological age= 71.04±10.45 years, 21.1% female, table 1). 11.4% patients were diagnosed with non-obstructive CAD with an atherosclerotic lumen reduction below 50% (chronological age= 70.60±9.38 years, 40.4% female), while a normal angiogram was observed in 17.5% of the participants (chronological age=71.04±10.45 years, 21.1% female). Participants diagnosed with obstructive CAD were more likely to be male (corrected p=3.88⨯10^−21^), chronologically and epigenetically older (corrected p=1.62⨯10^−8^ and corrected p=2.29⨯10^−6^ respectively) with an increased rDNAm age acceleration (corrected p=1.60⨯10^−9^) and lower HDL-C levels (corrected p=2.80⨯10^−5^) in comparison to patients with a normal angiogram. Systolic blood pressure, triglyceride level, estimated glomerular filtration rate (eGFR), diabetes mellitus type II status, lipoprotein (a) levels above 107 nmol/L (fifth percentile), BMI, physical activity according to the RAPA assessment, alcohol consumption and smoking behaviour were comparable between the above subcohorts. Clinical characteristics, including lifestyle and risk factors for CAD are listed in table 1.

**Table 1:**
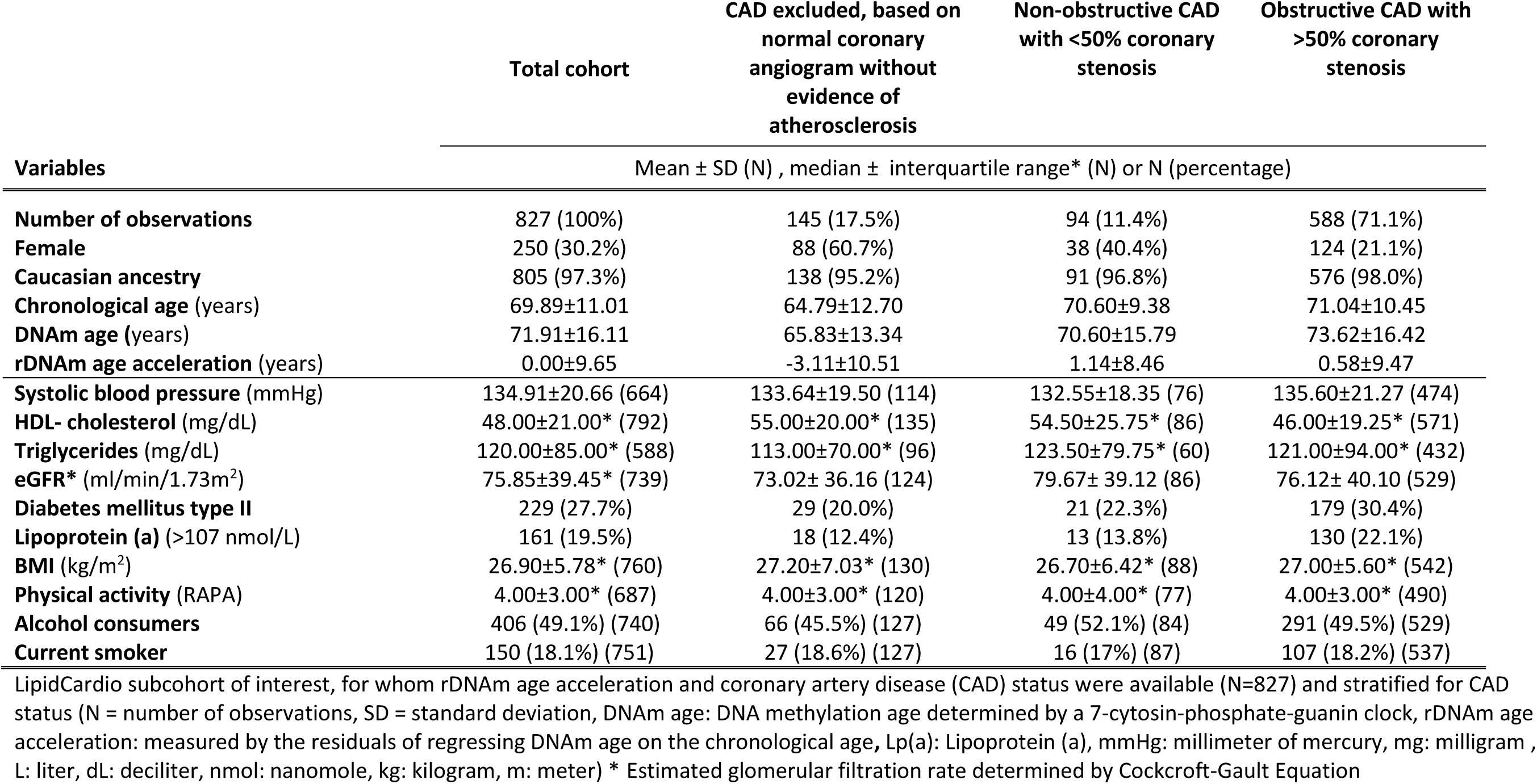
Descriptive statistics of LipidCardio subcohort of interest.

|  | Total cohort | CAD excluded, based on<br>normal coronary<br>angiogram without<br>evidence of<br>atherosclerosis | Non-obstructive CAD<br>with <50% coronary<br>stenosis | Obstructive CAD with<br>>50% coronary<br>stenosis |
| --- | --- | --- | --- | --- |
| Variables | Mean ± SD (N) , median ± interquartile range* (N) or N (percentage) |  |  |  |
| **Number of observations** | 827 (100%) | 145 (17.5%) | 94 (11.4%) | 588 (71.1%) |
| **Female** | 250 (30.2%) | 88 (60.7%) | 38 (40.4%) | 124 (21.1%) |
| **Caucasian ancestry** | 805 (97.3%) | 138 (95.2%) | 91 (96.8%) | 576 (98.0%) |
| **Chronological age (years)** | 69.89±11.01 | 64.79±12.70 | 70.60±9.38 | 71.04±10.45 |
| **DNAm age (years)** | 71.91±16.11 | 65.83±13.34 | 70.60±15.79 | 73.62±16.42 |
| **rDNAm age acceleration (years)** | 0.00±9.65 | -3.11±10.51 | 1.14±8.46 | 0.58±9.47 |
| **Systolic blood pressure (mmHg)** | 134.91±20.66 (664) | 133.64±19.50 (114) | 132.55±18.35 (76) | 135.60±21.27 (474) |
| **HDL- cholesterol (mg/dL)** | 48.00±21.00* (792) | 55.00±20.00* (135) | 54.50±25.75* (86) | 46.00±19.25* (571) |
| **Triglycerides (mg/dL)** | 120.00±85.00* (588) | 113.00±70.00* (96) | 123.50±79.75* (60) | 121.00±94.00* (432) |
| **eGFR\* (ml/min/1.73m<sup>2</sup>)** | 75.85±39.45* (739) | 73.02± 36.16 (124) | 79.67± 39.12 (86) | 76.12± 40.10 (529) |
| **Diabetes mellitus type II** | 229 (27.7%) | 29 (20.0%) | 21 (22.3%) | 179 (30.4%) |
| **Lipoprotein (a) (>107 nmol/L)** | 161 (19.5%) | 18 (12.4%) | 13 (13.8%) | 130 (22.1%) |
| **BMI (kg/m<sup>2</sup>)** | 26.90±5.78* (760) | 27.20±7.03* (130) | 26.70±6.42* (88) | 27.00±5.60* (542) |
| **Physical activity (RAPA)** | 4.00±3.00* (687) | 4.00±3.00* (120) | 4.00±4.00* (77) | 4.00±3.00* (490) |
| **Alcohol consumers** | 406 (49.1%) (740) | 66 (45.5%) (127) | 49 (52.1%) (84) | 291 (49.5%) (529) |
| **Current smoker** | 150 (18.1%) (751) | 27 (18.6%) (127) | 16 (17%) (87) | 107 (18.2%) (537) |

LipidCardio subcohort of interest, for whom rDNAm age acceleration and coronary artery disease (CAD) status were available (N=827) and stratified for CAD status (N = number of observations, SD = standard deviation, DNAm age: DNA methylation age determined by a 7-cytosin-phosphate-guanin clock, rDNAm age acceleration: measured by the residuals of regressing DNAm age on the chronological age, Lp(a): Lipoprotein (a), mmHg: millimeter of mercury, mg: milligram , L: liter, dL: deciliter, nmol: nanomole, kg: kilogram, m: meter) \* Estimated glomerular filtration rate determined by Cockcroft-Gault Equation

### rDNAm Age Acceleration and Cardio Vascular Disease Status

The mean rDNAm age acceleration (N=827, mean chronological age: 69.82±11.01 years, DNAm age: 71.91±16.11) was estimated 0.00±9.65 years by the 7-CpGs epigenetic clock trained in the BASE II cohort (Vetter et al., 2018).

The stratified analysis according to CAD status, showed that rDNAm age acceleration was significantly accelerated in patients diagnosed with non-obstructive CAD (N=94, rDNA age acceleration= 1.14±8.46, corrected p= 4.56⨯10^−4^) and patients diagnosed with obstructive CAD defined by >50% lumen reduction (N=588, rDNA age acceleration=0.58±9.47, corrected p= 2.05⨯10^−3^) compared to patients without any coronary artery stenosis a normal angiogram (N=145, rDNAm age acceleration= -3.11±10.51 years) (figure1). rDNAm age acceleration did not differ significantly between the groups of patients diagnosed with obstructive and non-obstructive CAD.

**Figure 1.**
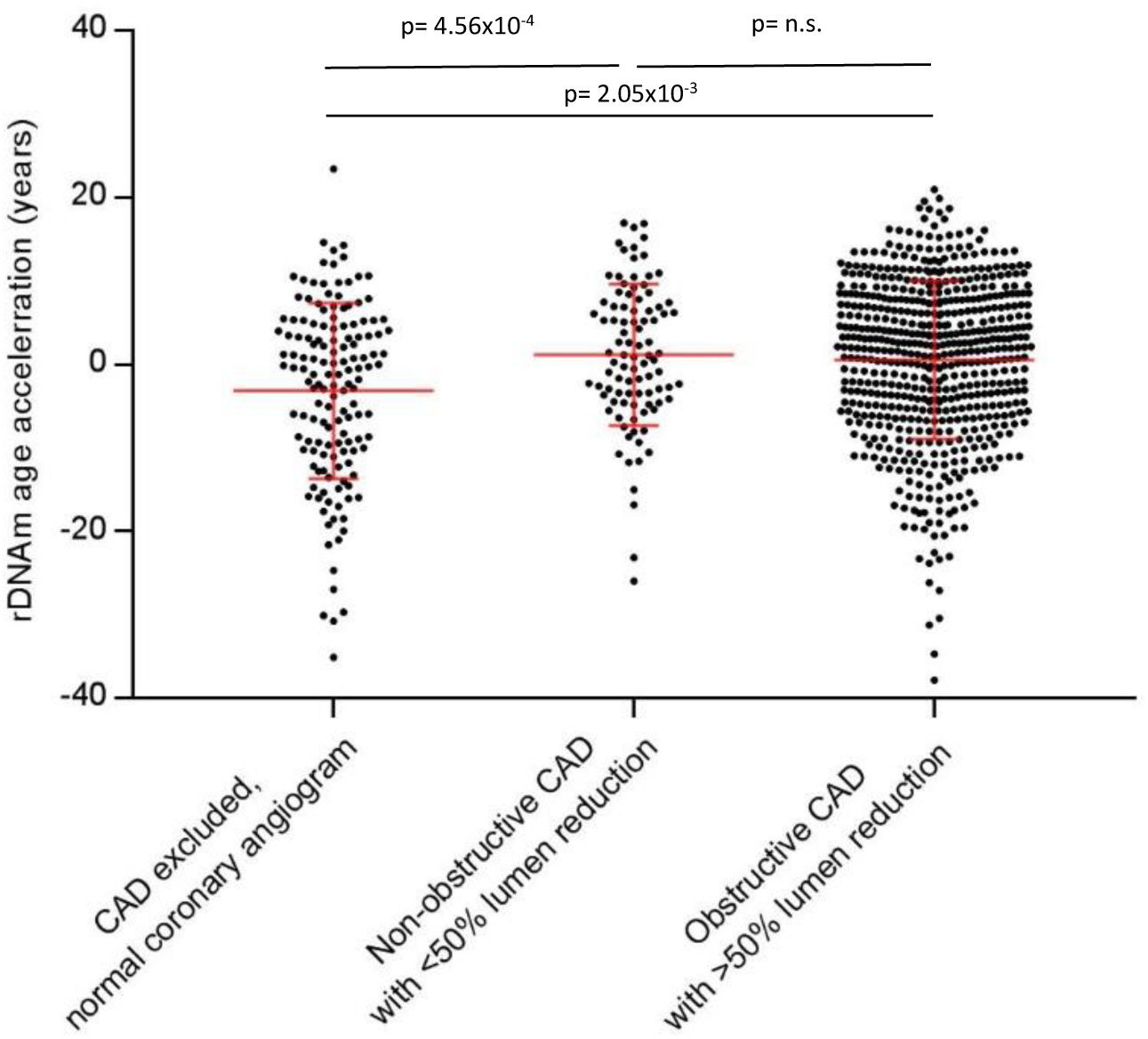
rDNAm age acceleration stratified according to coronary artery disease (CAD) status: (1) controls with normal angiogram (N=145, rDNAm age acceleration= -3.11±10.51 years), (2) non-obstructive arthrosclerosis (N=94, rDNA age acceleration= 1.14±8.46) and (3) obstructive CAD with a lumen reduction exceeding 50% (N=588, rDNA age acceleration=0.58±9.47), depicted are the mean and the standard deviation, p-values (p) are Bonferroni corrected (rDNAm age acceleration: residual DNA methylation age acceleration)

The association of rDNAm age acceleration and CAD status of subjects without coronary artery stenosis versus patients with obstructive CAD remained significant (ß=2.128, 95% CI 0.854 – 3.401, p= 1.12⨯10^−3^) after adjusting for systolic blood pressure, HDL-C, triglycerides levels, eGFR, Lipoprotein (a) levels, BMI, physical activity, alcohol and nicotine consumption by linear regression analysis (table 2). Moreover, rDNAm age acceleration was significantly associated with systolic blood pressure (ß=0.069, 95% CI 0.027 – 0.112, p= 1.44⨯10^−3^), sex (ß=-2.438, 95% CI -4.591 - -0.285, p= 2.65⨯10^−2^), eGFR (ß=0.040, 95% CI 0.011 – 0.069, p= 6.87⨯10^−9^) and smoking status (ß=-8.538, 95% CI -10.772 - -6.303, p= 2,45⨯10^−13^).

**Table 2.** Result of linear regression analysis.

| Dependent variable: rDNAm age acceleration |  |  |  |  |  |
| --- | --- | --- | --- | --- | --- |
| Covariable | $\beta$ | Standardized $\beta$ | Standard error | 95% CI | p-value |
| **CAD status** | **2.128** | **0.158** | **0.648** | **0.854 – 3.401** | **$1.12 \times 10^{-3}$** |
| **Systolic blood pressure (mmHg)** | **0.069** | **0.144** | **0.022** | **0.027 – 0.112** | **$1.44 \times 10^{-3}$** |
| **Sex** | **-2.438** | **0.113** | **1.095** | **-4.591 - -0.285** | **$2.65 \times 10^{-2}$** |
| **HDL- cholesterol (mg/dl)** | 0.037 | 0.064 | 0.031 | -0.024 - 0.098 | $2.23 \times 10^{-1}$ |
| **Triglycerides (mg/dl)** | -0.010 | -0.091 | 0.006 | -0.021 - 0.001 | $6.36 \times 10^{-2}$ |
| **eGFR (ml/min/1.73m<sup>2</sup>)** | **0.040** | **0.122** | **0.015** | **0.011 – 0.069** | **$6.87 \times 10^{-9}$** |
| **Diabetes mellitus type II** | -1.671 | -0.075 | 1.057 | -3.750 – 0.407 | $1.15 \times 10^{-1}$ |
| **Lipoprotein (a) >107 nmol/L** | 0.262 | 0.011 | 1.122 | -1.944 – 2.467 | $8.15 \times 10^{-1}$ |
| **BMI (kg/m<sup>2</sup>)** | -0.043 | 0.020 | 0.101 | -0.241 – 0.155 | $6.69 \times 10^{-1}$ |
| **Physical activity (RAPA)** | -0.264 | -0.053 | 0.228 | -0.712 – 0.184 | $2.45 \times 10^{-1}$ |
| **Alcohol consumption (yes/no)** | -1.466 | -0.073 | 0.944 | -3.322 – 0.391 | $1.21 \times 10^{-1}$ |
| **Smoking (yes/no)** | **-8.538** | **-0.346** | **1.137** | **-10.772 - -6.303** | **$2.45 \times 10^{-13}$** |
| **R<sup>2</sup>= 0.232 , corrected R<sup>2</sup>= 0.209 , F= 9.825** |  |  |  |  |  |

Result of linear regression analysis, regressing coronary artery disease (CAD) status and its risk factors on the rDNAm age acceleration (rDNAm age acceleration: measured by the residuals of regressing DNAm age determined by a 7-cytosin-phosphate-guanin clock on the chronological age, CI: confidence interval, mmHg: millimeter of mercury, mg = milligram, L = liter, dL = deciliter, nmol: nanomole, kg= kilogram, m: meter)

### Association of cardiovascular disease status and the methylation status at the seven cytosin-phosphate-guanine nucleotides of interest

Cg16386080 (odds ratio (OR) =9.897, 95% CI 1.188 – 82.472, p=0.034), cg02228185 (OR=0.182, 95% CI 0.036 – 0.913, p=0.038) and cg19761273 (OR=1.29⨯10^−4^, 95% CI 5.922⨯10^−7^ – 0.28, p=0.001) were significantly associated with the CAD status (CAD negative vs. obstructive CAD) (table 3).

**Table 3.** Result of multiple logistic regression.

**Dependent variable: CAD status** (patients with normal angiogram vs. patients with obstructive CAD)
| Covariable | Odds ratio | 95% CI | p-value |
| --- | --- | --- | --- |
| **Cg09809672 (A)** | 1.311 | 0.203 – 8.465 | 0.776 |
| **Cg24768561 (B)** | 1.959 | 0.196 – 19.619 | 0.567 |
| **Cg16386080 (C)** | **9.897** | **1.188 – 82.472** | **0.034** |
| **Cg02228185 (E)** | **0.182** | **0.036 – 0.913** | **0.038** |
| **Cg25809905 (F)** | 0.679 | 0.083 – 5.045 | 0.679 |
| **Cg19761273 (G)** | **1.29x10<sup>-4</sup>** | **5.922x10<sup>-7</sup> – 0.28** | **0.001** |
| **Cg17471102 (H)** | 0.129 | 0.007 – 2.474 | 0.174 |
| **Nagelkerke's R<sup>2</sup>= 0.089, n=733** |  |  |  |

Result of multiple logistic regression, regressing the methylation status at the seven cytosine-phosphate-guanine nucleotides of interest individually on the cardiovascular disease (CAD) status (CAD negative vs. obstructive CAD) (CI: confidence interval)

### Literature review

While we identified five original research articles studying the epigenetic clock in the context of CVD and CAD (P. Dugué et al., 2018; Horvath, Gurven, Levine, Trumble, Kaplan, Allayee, Ritz, Chen, Lu, Rickabaugh, Jamieson, Sun, Li, Chen, Quintana-Murci, et al., 2016; Lind et al., 2017; Perna et al., 2016; Roetker et al., 2018), further studies have researched a single or multiple CVD and CAD risk factors (Supplementary Table 1). To our knowledge we are first to investigate the epigenetic clock with respect to lipoprotein (a).

## Discussion

The epigenetic clock variable DNAm age acceleration is currently being evaluated as a biomarker of aging. We have shown previously that DNAm age (measured by a seven-CpGs epigenetic clock trained in the LipidCardio cohort) is correlated with chronological age, independent of the established biomarker of aging - relative telomere length - in the LipidCardio cohort (Banszerus et al., 2019), we here aimed to validate a seven-CpGs epigenetic clock’s potential (trained in the population-based and mostly healthy Berlin Aging Study II cohort (Vetter et al., 2018)) to predict premature biological aging in patients diagnosed with CAD.

Suspected CAD was confirmed in 71.1% (N=588) of the LipidCardio participants (for whom information on rDNAm age acceleration were available), based on the result of the coronary angiography. While 11.4% (N=95) patients were diagnosed with non-obstructive atherosclerosis with a lumen reduction below 50%, 17.5% (N=145) were allocated to the group with no evidence of atherosclerosis confirmed based on a normal angiogram. Participants diagnosed with obstructive CAD were more likely to be male (corrected p=3.88⨯10^−21^) and of advanced chronological and epigenetic age (corrected p=1.6⨯10^−8^ and corrected p=2.29⨯10^−6^ respectively) compared to patients with a normal angiogram. rDNAm age was significantly accelerated in patients diagnosed with non-obstructive CAD (rDNA age acceleration= 1.14±8.46, corrected p= 4.56⨯10^−4^) and obstructive CAD (rDNA age acceleration=0.58±9.47, corrected p= 2.05⨯10^−3^), compared to patients with a normal angiogram. rDNAm age acceleration was not significantly different between patients with non-obstructive CAD with a lumen reduction <50% and patients with obstructive CAD with a lumen reduction >50%. The association of CAD status (normal angiogram versus obstructive CAD) remained significant after adjusting for known co-morbidities and clinical risk factors, including systolic blood pressure, HDL-C, triglyceride levels, diabetes mellitus II status, eGFR, lipoprotein (a), BMI, including physical activity assessed by the RAPA questionnaire, alcohol and nicotine consumption.

While previous research has focused on evaluating Horvath or Hannum’s epigenetic clocks to study CVD and CAD, yielding inconclusive results, we are to our knowledge first to report an association of CAD and rDNAm age acceleration, estimated by a reduced seven-CpGs epigenetic clock. Further we report a significant association of the rDNAm age acceleration with the CAD risk factors including systolic blood pressure (ß=0.069, 95% CI 0.027 – 0.112, p= 1.44⨯10^−3^), sex (ß=-2.438, 95% CI -4.591 - -0.285, p= 2.65⨯10^−2^), eGFR (ß=0.040, 95% CI 0.011 – 0.069, p= 6.87⨯10^−9^) and smoking (ß=-8.538, 95% CI -10.772 - -6.303, p= 2,45⨯10^−13^). HDL-cholesterol levels, triglyceride levels, diabetes mellitus type II status, lipoprotein (a) levels, BMI, physical activity and alcohol consumption were not associated with rDNAm age acceleration in the LipidCardio cohort.

### S*ystolic blood pressure*

The importance of s*ystolic blood pressure* as a risk factor for CAD is emphasized by studies like the meta-analysis by Ettehad et al. including 613,815 from 123 studies, reporting that every 10 mmHg reduction in systolic blood pressure, reduces the risk of CAD significantly (relative risk: 0.83, 95% CI 0.78-0.88) (Ettehad et al., 2016). In the LipidCardio cohort, we observed a positive association of rDNAm age acceleration and systolic blood pressure (ß=0.069, 95% CI 0.027 – 0.112, p= 1.44⨯10^−3^). Consistent with the above finding, Lind et al. reported significant positive association (ß=0.02, 95% CI 0.00 - 0.03, p= 0.012) of the DNAm age acceleration, determined by the difference of the Horvath estimate and the chronological age, and systolic blood pressure in the PIVOUS study (Lind et al., 2017).

Interestingly, McCartney et al., who studied increased blood pressure in the context of Alzheimer’s disease, observed a positive association of the extrinsic Hannum DNA age acceleration (ß=0.177, 95% CI 0.064 – 0.29, p=0.002), but not the intrinsic Horvath rDNA age acceleration in the Generation Scotland study (N=5100, chronological age: 48.51±13.99 years, 38.4% female) (McCartney et al., 2018).

### Sex

While CAD has been perceived as a man’s disease in the past, contributed by the different incident rates before the fifth decade with a late-onset rapid increase and a predominantly atypical clinical presentation in females, the importance of biological sex as a risk factor of CAD is being recognized progressively (Pathak, Shirodkar, Ruparelia, & Rajebahadur, 2017). In the LipidCardio cohort, the percentage of males, affected by CAD was greater than females. Moreover, female sex was associated with lower rDNAm age acceleration in the cohort, independent of the CAD status (ß=-2.438, 95% CI -4.591 - -0.285, p= 2.65⨯10^−2^). The diversion of male and female DNAm age acceleration has been studied in a range of cohorts, supporting the hypothesis that male aging is accelerated compared to females. For instance, Roetker et al. reported a 0.7 – 1.5 years higher average DNAm age acceleration in male participants compared to females in the 2,543 participants of the ARIC study (Roetker et al., 2018). Supporting evidence of a positive association of male sex and accelerated residual epigenetic aging was also observed by Dugué et al. in the Melbourne Collaborative Cohort Study (Hannum’s intrinsic age acceleration: ß=1.37 years, p<0.001 and Horvath’s extrinsic acceleration: ß=1.23 years, p=0.005) as well as by Zhao et al. in the in the Generic Epidemiology Network of Arteriopathy (GENOA) study (N=1100, chronological age= 57.05±10.48 years, 29% female) (Horvath’s intrinsic age acceleration: ß=1.038 years, p=0.003 and Hannum’s extrinsic acceleration: ß=2.405 years, p=1.94⨯10^−8^) (P. A. Dugué et al., 2018; Zhao et al., 2019).

### HDL-cholesterol and triglyceride levels

Low HDL-cholesterol and high triglyceride levels have independently been associated with CAD (Lee et al., 2017). In the LipidCardio cohort, both CAD risk factors were not associated with rDNAm age acceleration. Similarly previous studies have found either a weak or no association: Horvath et al., who observed no association of HDL-cholesterol levels and intrinsic or extrinsic age acceleration, reported a weak association of triglyceride levels and extrinsic age acceleration (ß=0.004, p=0.04), but not intrinsic age acceleration (both measured based on the Horvath clock) in the WHI cohort (Horvath, Gurven, Levine, Trumble, Kaplan, Allayee, Ritz, Chen, Lu, Rickabaugh, Jamieson, Sun, Li, Chen, Quintana-Murci, et al., 2016). A second study by Lind et al. found neither evidence of an association of the DNAm age acceleration (determined by the difference of the Horvath estimate and the chronological age) and serum triglyceride levels nor HDL-cholesterol levels in the PIVOUS cohort (Lind et al., 2017). Irvine et al. detected a marginal association of the Horvath DNAm age acceleration and postprandial HLD-cholesterol (ß=0.05±0.03,p=0.05) in 830 participants of the Generics pf Lipid Lowering Drugs and diet Network (GOLDN) study (N=830, middle-age, ∼ 50% females) (Irvin et al., 2018). The Hannum DNAm age acceleration was inversely associated with fasting HDL-cholesterol levels (ß=-0.11±0.04, p=0.02) and positively associated with postprandial triglyceride levels (ß=0.004±0.001, p=0.02) (Irvin et al., 2018).

Roetker et al. reported a weak negative association (ß=-0.65p=0.04) of HDL cholesterol and Horvath DNAm age acceleration, but not the Hannum age acceleration in the ARIC study (Roetker et al., 2018). Interestingly, a niche clinical study accessing epigenetic age of the placenta in the context of maternal dyslipidemia in early pregnancy by Shrestha et al. found evidence of an association of low maternal HDL-cholesterol levels (but not high triglyceride levels) and accelerated aging of the placenta (assessed placental age acceleration by the difference of a 62-CpGs estimate and the gestational age) of normal weight mothers and female offspring in 262 women participating in the Fetal Growth Studies - Singleton cohort (Shrestha et al., 2019). While the above findings are considerable, its comparability to our findings is limited.

### Estimated glomerular filtration rate (eGFR)

Moderately reduced eGFR of 60-89 mL/min per 1.73 m^2^, as well as chronical kidney disease, defined by a eGFR <60 mL/min per 1.73 m^2^ have been associated whit the cardiometabolic risk factors (including hypertension, diabetes mellitus, obesity and dyslipidemia) and a significantly higher risk of atherosclerotic CVD and CAD (Lu et al., 2016). In the LipidCardio cohort, a median clearance rate of 75.85±39.45 ml/min/1.73m^2^ indicated reduced kidney function in the total cohort. Moreover, eGFR and rDNAm age acceleration were associated positively in the study cohort (ß=0.040, 95% CI 0.011 – 0.069, p= 6.87⨯10^−9^). Contrarily, Horvath et al. observed a significant association of creatinine levels and extrinsic age acceleration (ß=1.985, p=0.008), but no intrinsic age acceleration in the WHI cohort (Horvath, Gurven, Levine, Trumble, Kaplan, Allayee, Ritz, Chen, Lu, Rickabaugh, Jamieson, Sun, Li, Chen, Quintana-Murci, et al., 2016).

### Diabetes mellitus type II

Diabetes mellitus type II is associated with an increased risk of CAD (Teven et al., 1998). Indeed 22.3% participants diagnosed with non-obstructive CAD and 30.4% of participants diagnosed with obstructive CAD, had previously been diagnosed with diabetes mellitus type II in the LipidCardio cohort. Nonetheless, diabetes mellitus type II status was not associated with rDNAm age acceleration.

Agreeing with the above, an association with diabetes mellitus type II status (yes/no) was neither detected by the intrinsic Horvath estimate or the extrinsic Hannum estimate of epigenetic aging assessed in the Generation Scotland Study (McCartney et al., 2018), nor by intrinsic or extrinsic Horvath age acceleration in the WHI cohort (Horvath, Gurven, Levine, Trumble, Kaplan, Allayee, Ritz, Chen, Lu, Rickabaugh, Jamieson, Sun, Li, Chen, Quintana-murci, et al., 2016). Moreover, first-line diabetes mellitus type II treatment with metformin did not decelerate extrinsic or intrinsic epigenetic aging (measured by the Hannum or Horvath clock respectively) in the 4,173 female participants of the WHI (Quach, Levine, Tanaka, Lu, Chen, Ferrucci, et al., 2017). Only Dugué et al. showed a positive association of intrinsic DNAm age acceleration, measured by the Horvath estimate (but not the extrinsic Hannum estimate) and diabetes mellitus type II status (yes/no) in the Melbourne Collaborative Cohort Study (ß=1.46 years, p=0.04) (P. A. Dugué et al., 2018).

### Lipoprotein (a)

Due to its atherogenic properties, elevated lipoprotein (a) levels have been identified as risk factor of CAD (Kronenberg, 2016). To our knowledge, lipoprotein (a) levels have not been studied in the context of epigenetic aging measured by an epigenetic clock previously. In the LipidCardio cohort we found no evidence of an association of elevated lipoprotein (a) levels (defined by the fifth quintile of >107 nmol/L) and rDNAm age acceleration.

### BMI

A systematic review and meta-analysis by Ryan et al. recently revised existing literature reporting on the epigenetic clock’s association with lifestyle factors including BMI as well as alcohol consumption and smoking (Ryan et al., 2019), concluding that there is an association between BMI and epigenetic ageing. (Ryan et al., 2019).

Moreover, three large scale studies have provided supporting evidence of an association previously: In the Generation Scotland study, including 5100 participants, intrinsic (ß=0.089, 95% CI 0.06-0.11, p<0.001) and extrinsic (ß=0.061, 95% CI 0.03-0.087, p<0.001) epigenetic aging, measured by the Horvath or Hannum estimate respectively, were both associated with BMI (McCartney et al., 2018). Quach et al. reported a weak association of intrinsic (r=0.08, p=1⨯10^−6^) and extrinsic (r=0.09, p=2⨯10^−8^) epigenetic aging, measured by the Hannum or Horvath estimate respectively, with BMI, in the 4142 participants of the WHI and the InChIANTI cohort (Quach, Levine, Tanaka, Lu, Chen, Ferrucci, et al., 2017). In the Melbourne Collaborative Cohort Study (overweight participants with BMI 25-30 (ß=0.69 years, p=0.01), obsessed participants with BMI 30-35 (ß=1.28 years, p=0.001) and participants with BMI >35 (ß=1.70 years, p=0.01) showed significantly accelerated intrinsic Hannum epigenetic aging, compared to lean individuals with a BMI<25 (P. A. Dugué et al., 2018).

In the LipidCardio cohort we found no evidence of an association of BMI (N=760, median=26.90±5.78) and rDNAm age acceleration measured by the seven-CpG epigenetic clock. Which is in line with Zhao et al., who found no evidence of an association of Hannum’s extrinsic or Horvath’s intrinsic residual epigenetic age acceleration with BMI in the GENOA study (Zhao et al., 2019).

### Physical activity

Regular physical activity has been suggested to prevent and overt atherosclerosis and CAD (Linke, Erbs, & Hambrecht, 2008). In the LipidCardio cohort, we found no association of rDNAm age acceleration and the RAPA scores, assessing physical activity (N=687, median=4.00±3.00). This is in line with an increasing body of evidence suggesting an (at most) minor immediate effect of an active lifestyle on the epigenetic aging estimates.

Quach et al. studied 4142 participants of the WHI and the InChIANTI cohort and found no association between extrinsic or intrinsic DNAm age acceleration (measured by the Hannum or Horvath clock respectively) and sedentary or active lifestyle in previous years (Quach, Levine, Tanaka, Lu, Chen, Ferrucci, et al., 2017). In the Melbourne Collaborative Cohort Study, no association was detected between intrinsic age acceleration measured by the Hannum estimate (ß=0.18 1.08, SE 0.11, p=0.11) or the Horvath estimate (ß=0.07 1.08, SE 0.13, p=0.59) and physical activity, evaluated by a four point score (P. Dugué et al., 2018). In the Lothian Birth Cohort 1936 (N=248, chronological age= 79 years) intrinsic epigenetic age acceleration and extrinsic age acceleration (defined by the residuals of Horvath or Hannum estimate regressed on chronological age respectively), were no longer significantly associated with reduced step count and sit-to-standing transitions, after adjusting for multiple testing (Gale et al., 2018). Comparably, a smaller study including 100 black US residents (Family and Community Health Study (FACHS), chronological age= 48.5±9.2 years) found neither evidence of an association of accelerated ageing (determined by the regression of the Horvath estimate on chronological age) and BMI, nor exercise, defined by the number of physical activity lasting 30 minutes with the past seven days (Simons et al., 2017). Further, discordant leisure-time physical activity did not coincide with a difference in the Horvath age estimate (mean chronological age=60.4 years, DNAm age= 60.7 and 61.8 years respectively) in seven monozygotic and nine dizygotic twins, who were follow-up over a period of 32 years (Sillanpää et al., 2019).

### Alcohol consumption

Moderate alcohol consumption has been suggested to be CAD-protective, whereas instable drinking behaviour has been linked with an increased risk for incidence of CAD (O’Neill et al., 2018). In accordance with the above, Beach et al. reported on a dose-dependent effect of alcohol consumption on the Hannum age estimated in 836 participants of a Caucasian/Hispanic cohort previously studied by Hannum et al. (Hannum et al., 2013) and the FACHS cohort (N=656, chronological age: 63.4 ± 14.8 years and N=180, chronological age: 48.9 ± 8.6 years, 61.7% female respectively) with accelerated epigenetic aging in low and heavy drinkers and decelerated aging in moderate consumers (Beach et al., 2015). Consistently, Quach et al. reported a negative association of extrinsic (r=-0.07, p=3⨯10^−5^) but not intrinsic epigenetic aging (measured by the Horvath or Hannum estimate respectively), with moderate alcohol consumption, in a combined cohort of 4,575 participants of the WHI and the InChIANTI study (Quach, Levine, Tanaka, Lu, Chen, Ferrucci, et al., 2017). Rosen et al. suggested a tissue-dependent association of alcohol dependence and Horvath rDNAm age acceleration, based on the study of blood samples (N=129, p=1⨯10^−4^), liver samples (N=92, p=6.9⨯10^−3^) and prefrontal cortex samples (n=46, p=n.s.) (Rosen et al., 2018).

Further, a clock-dependent association has been suggested, due to inconsistent findings in other studies: In the GENOA study, alcohol consumption was associated with higher 1030-CpGs-Grim-age acceleration (p = 2.76 × 10−9) and 513-CpGs-Pheno age acceleration (p = 0.038), but neither extrinsic Hannum rDNAm age acceleration nor intrinsic Horvath rDNAm age acceleration (Zhao et al., 2019). Moreover, Luo et al. studied 532 participants (chronological age: 40.7±12.5 years, 44.3% female) and observed a 1.38-year rDNAm age acceleration (p=0.02), according to the Levine clock, adjusted for blood cell count, sex, race, BMI and smoking behavior in heavy, chronic alcohol consumers, but no association with the Horvath or Hannum clock (M. Levine et al., 2018; Luo et al., 2019). Consistently, Horvath et al. found no evidence of an association of alcohol drinking, stratified according to never, light, moderate and heavy consumption and IEAA or EEAA in the 1462 postmenopausal female participants of the WHI study (Horvath, Gurven, Levine, Trumble, Kaplan, Allayee, Ritz, Chen, Lu, Rickabaugh, Jamieson, Sun, Li, Chen, Quintana-murci, et al., 2016). No association of low, or high alcohol intake and intrinsic age acceleration determined by Hannum and Horvath clocks was reported in the Melbourne Collaborative Cohort Study (P. A. Dugué et al., 2018). A smaller study, including hundred female US-American residence (chronological age: 48.5±9.2 years) found no evidence of an association of alcohol consumption, assessed as one of multiple indicators of health-related behavior, and the Hannum age acceleration (Simons et al., 2017). While, 49.1% (N=740) of the LipidCardio participants reported to consume alcohol, alcohol consumption and rDNAm age acceleration measured by a seven-CpGs epigenetic clock were not associated significantly in the LipidCardio cohort.

### Smoking

Even moderate tobacco consumption (one cigarette a day) is associated with a significant increase in the risk for heart disease (Hackshaw, Morris, Boniface, Tang, & Milenkovi, 2018). In the LipidCardio cohort 18.1% (N=751) of the participants reported to be current smokers. Surprisingly, smoking was associated with a decrease in rDNAm age acceleration (ß=-8.538, 95% CI -10.772 - -6.303, p= 2,45⨯10^−13^).

A number of studies have assessed the association of DNAm age acceleration and smoking, concluding no association (N=16,844 participants, included in a review by Ryan et al. (Ryan et al., 2019)) (Binder et al., 2018; Gao, Zhang, Breitling, & Brenner, 2016; Horvath, Gurven, Levine, Trumble, Kaplan, Allayee, Ritz, Chen, Lu, Rickabaugh, Jamieson, Sun, Li, Chen, Quintana-Murci, et al., 2016; Irvin et al., 2018; M. E. Levine et al., 2015; Quach, Levine, Tanaka, Lu, Chen, Ritz, et al., 2017; Simons et al., 2017). In agreement with the above studies, Simkin et al. found no association of smoking status and age acceleration (determined by the difference of the Horvath estimate and chronological age) in blood samples from 152 participants of the MRC National Survey of Health and Development (NSHD) study, while age acceleration was significantly reduced in buccal cells retrieved from current smokers (N=790, chronological age: 53.44±0.16 years, 21% current smoker, p=0.001) (Simpkin et al., 2017).

Contrarily, a positive dose-dependent association of cigarette consumption and Hannum DNAm age acceleration (ß=-0.8730, p<0.001) was reported in the cohort assessed by Beach et al. (introduced above) (Beach et al., 2015). Moreover, McCartney et al. observed a positive association of smoking in pack years the extrinsic Hannum DNA age acceleration (ß=0.059, 95% CI 0.033 – 0.086, p<0.001), but not the intrinsic Horvath rDNA age acceleration in the Generation Scotland study (McCartney et al., 2018). In the Melbourne Collaborative Cohort Study, both Hannum’s intrinsic age acceleration and Horvath’s extrinsic acceleration were associated positively with former smoking <15 years prior (N=533, ß=0.65±0.33, p=0.05 and ß=0.78±0.3, p=0.03 respectively) and current smoking (<20 cigarettes/day, N=159, ß=2.12±0.52, p<0.001 and ß=1.33±0.58, p=0.02 respectively) (P. A. Dugué et al., 2018). Interestingly, infants of smoking mothers, had a higher risk of a positive age acceleration, measured by the difference of the Horvath age estimate and chronological age in cord-blood samples, compared to infants with non-smoking mothers (N=80, OR= 3.17, 95% CI 1.05-9.56) (Javed et al., 2016).

In the GENOA study, a dose-dependent effect on Grim-age acceleration and Pheno-age acceleration was observed in former smokers (ß=2.37, p<0.001 and ß=1.37, p=0.015 respectively) and current smokers (ß=7.68, p<0.001 and 2.15, p=0.001 respectively), but not the intrinsic Horvath rDNAm age acceleration (Zhao et al., 2019). Extrinsic Hannum rDNAm age acceleration was significantly associated with former smoking (ß=0.95, p<0.009), but not current smoking status (Zhao et al., 2019).

### CpGs and CAD status

As the methylation status at cg16386080 increases, the odds for obstructive CAD increases approximately by a factor 10, while the odds for obstructive CAD decrease with increasing methylation status at cg02228185 and cg19761273.

### Limitations

The predictive power of the seven CpG epigenetic clock might have been undermined by comparing patients diagnosed with CAD and patients with a susceptibility for CAD, for whom CAD was excluded based on a normal angiogram rather than healthy controls.

As mentioned previously elsewhere (Banszerus et al., 2019), the leucocyte distribution undergoes a fluctuation during the course of aging. Due to a lack of data on the leukocyte distribution of the LipidCardio participants, we chose an established method to determine rDNAm age, without correcting for the leukocyte distribution, which needs to be considered, when interpreting our results. However, different methods to determine DNAm with and without correcting for the blood cell count have yielded no significant difference and were rated as negligible (Banszerus et al., 2019).

Besides the seven-CpGs employed here has only been applied in the BASE-II previously, consequently the comparability with findings yielded based on other epigenetic clocks (like Horvath and Hannum) is limited. Complicated by a multitude of clocks employed, the inconsistent definition of DNAm age acceleration and the remarkable inconstancy of findings across different tissues and study populations, the comparability between studies is limited.

## Conclusion

Despite the increasing number of studies assessing CVD, CAD and its risk factors identified and discussed here, robust evidence to validate DNAm age acceleration as a biomarker of aging is scarce, due to remarkably inconsistent findings across studies - even when the same clocks have been chosen. While the seven-CpGs epigenetic clock employed here was able to monitor premature aging in CAD patients compared to patients with normal angiogram, its ability to predict accelerated aging with respect to CVD risk factors was limited to systolic blood pressure, sex, eGFR and potentially smoking. Whether the seven-CpGs epigenetic clock is applicable to assess biological aging in other cohorts and across further ageing-associated phenotypes, remains to be explored further.

## Data Availability

Due to concerns for participant privacy, data are available only upon request. External scientists may apply to the Steering Committee of LipidCardio for data access. Please contact the study coordinating PI Ilja Demuth at ilja.demuth@charite.

## Acknowledgments

We would like to acknowledge Gabriele Hildebrand and Susanne Rothe for their technical advice on the conduction of the laboratory assays. Further, we would like to thank the patients of the Department of Cardiology at the Charité – Universitätsmedizin Berlin, who were willing to participate in the study.

## Supplementary Figure 1 & Supplementary Table 1

**Supplementary Figure 1:**
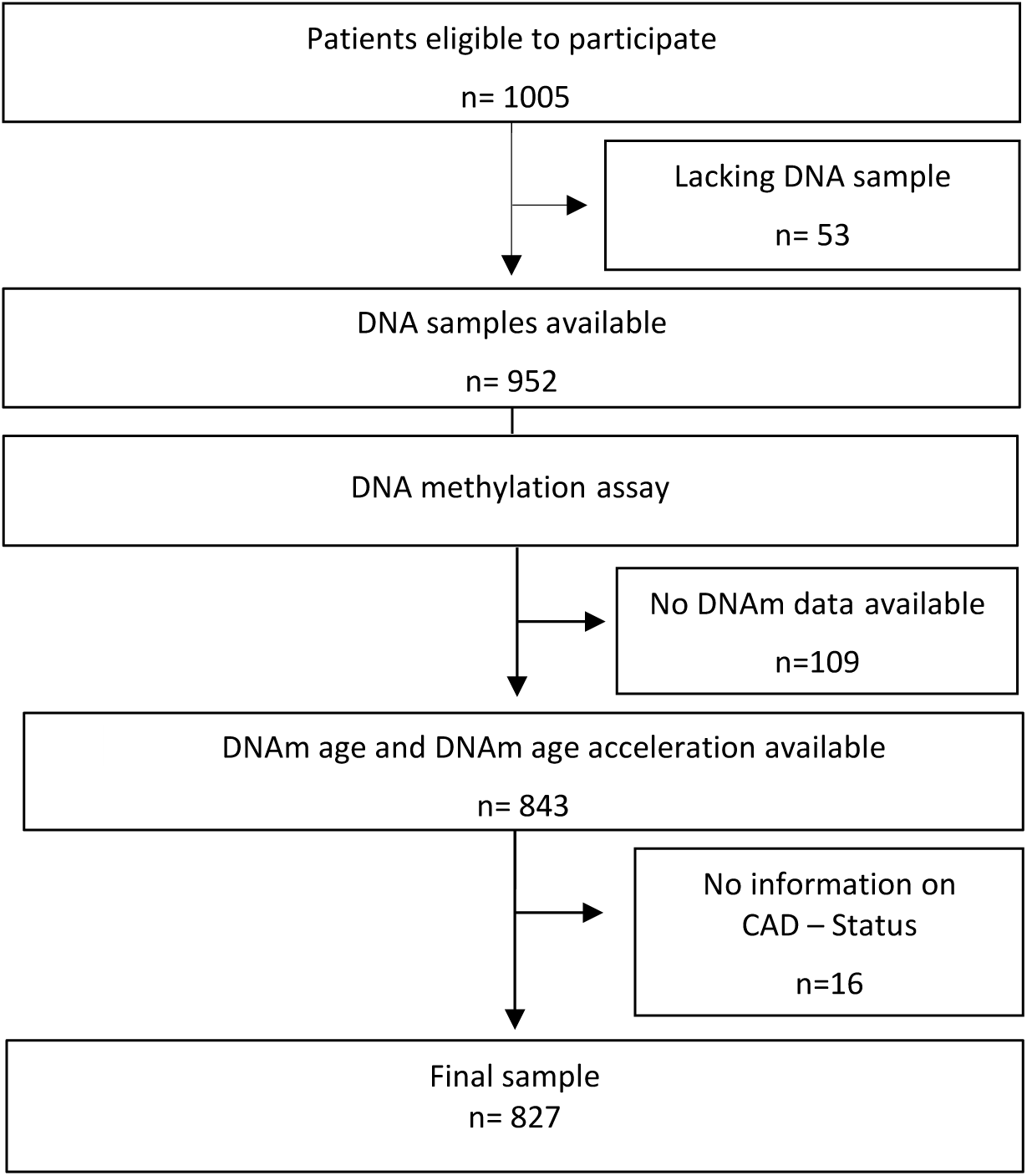
Flow chart – final sample.

**Supplementary Table 1:**
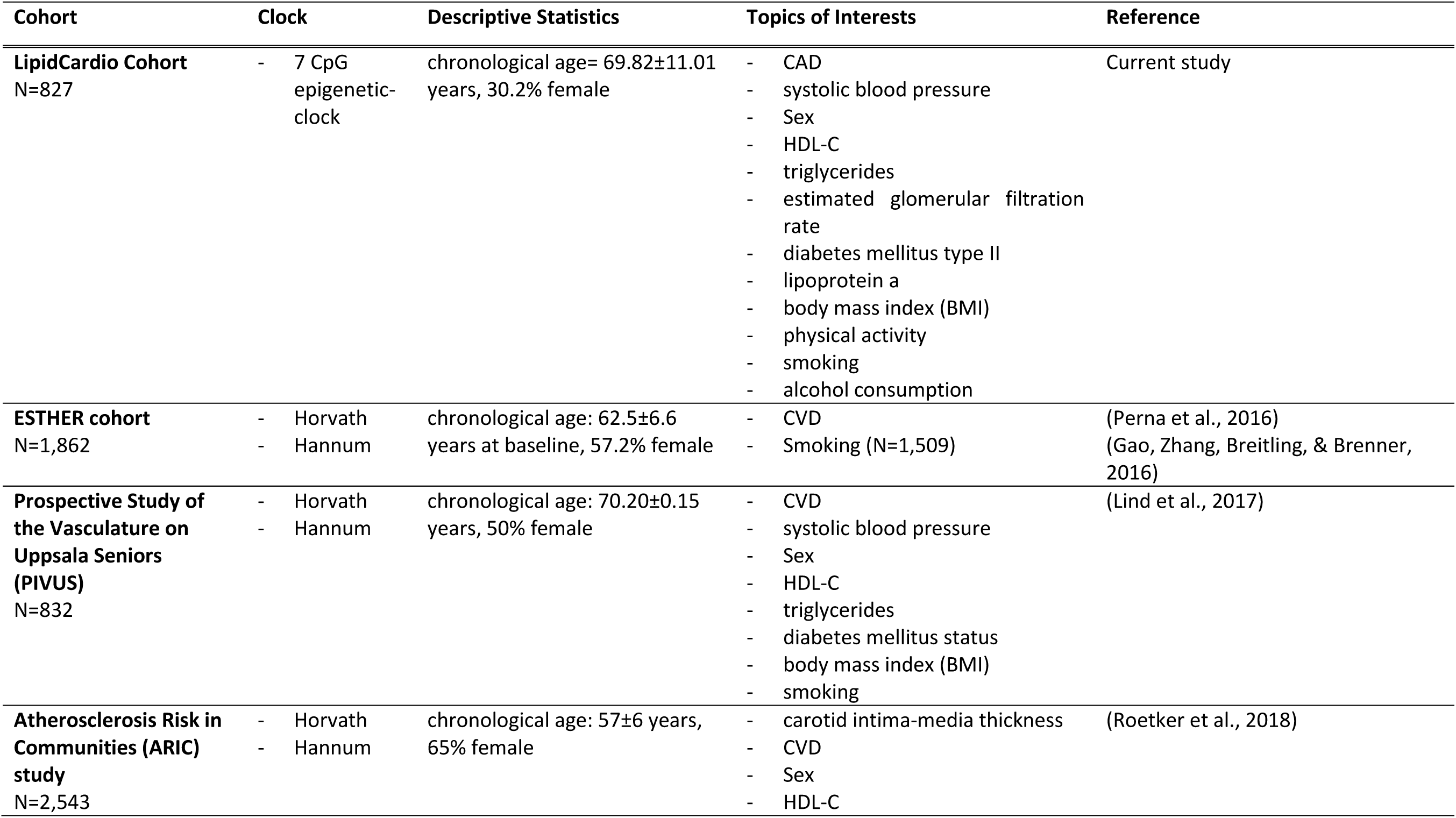
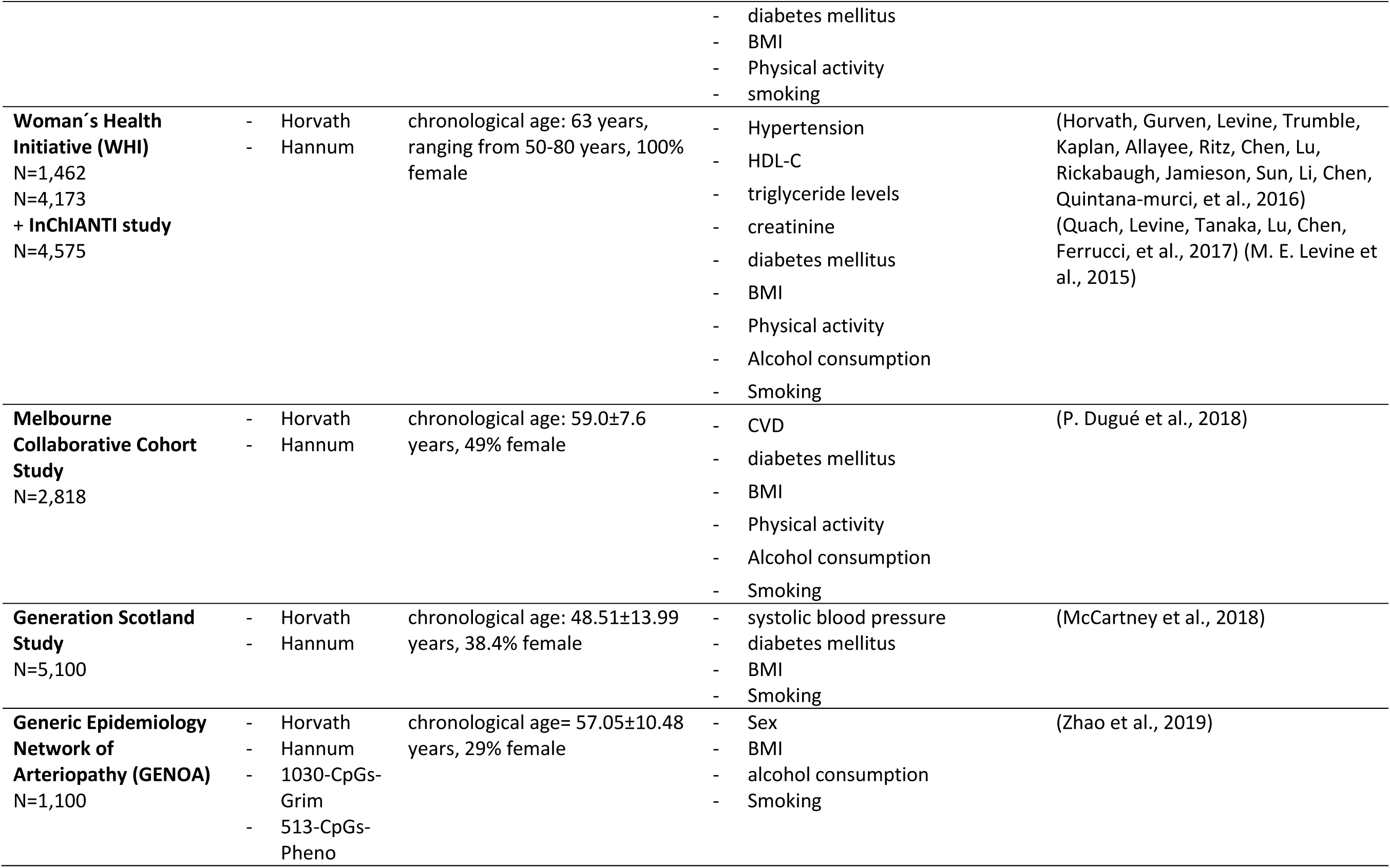
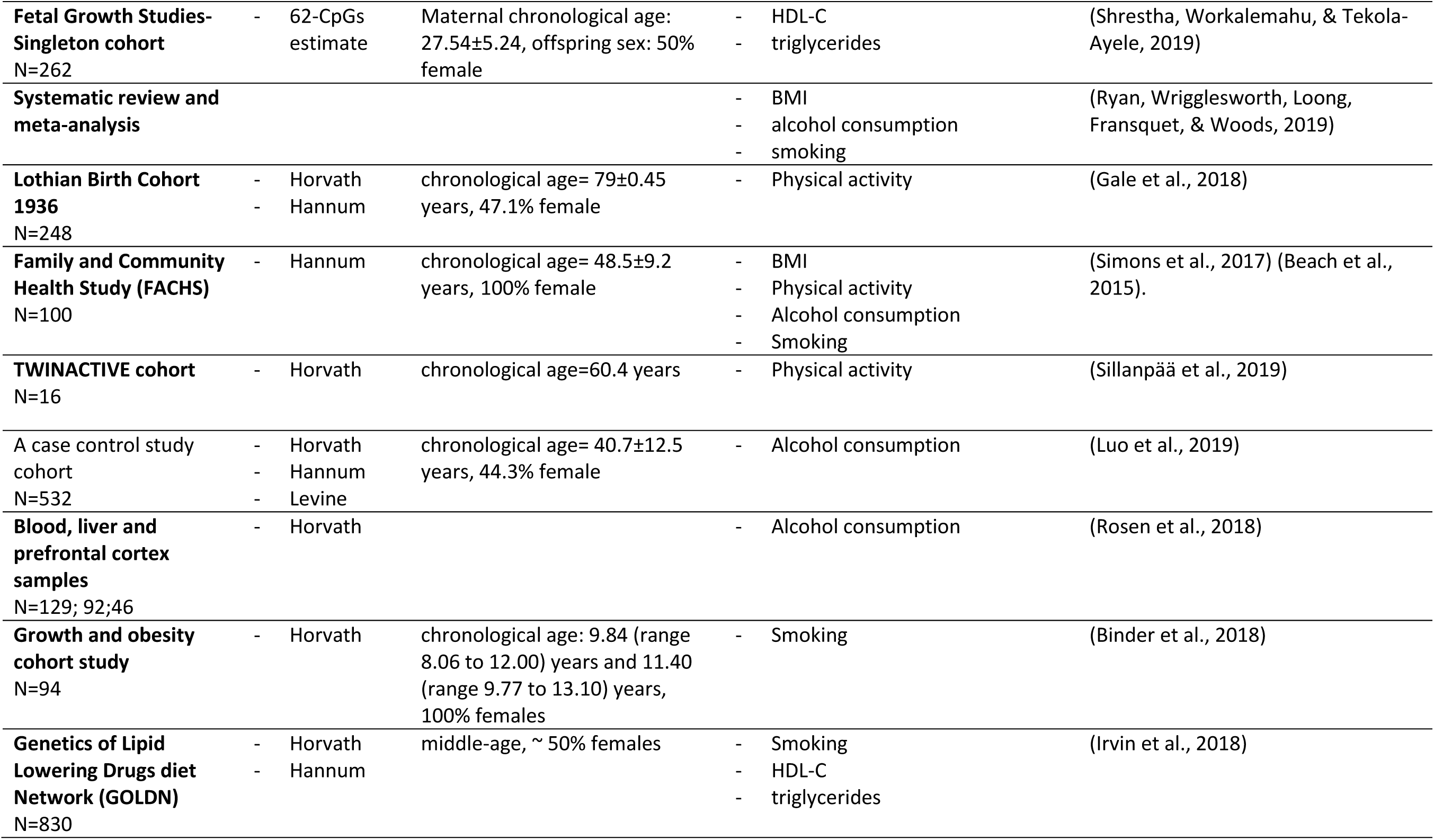
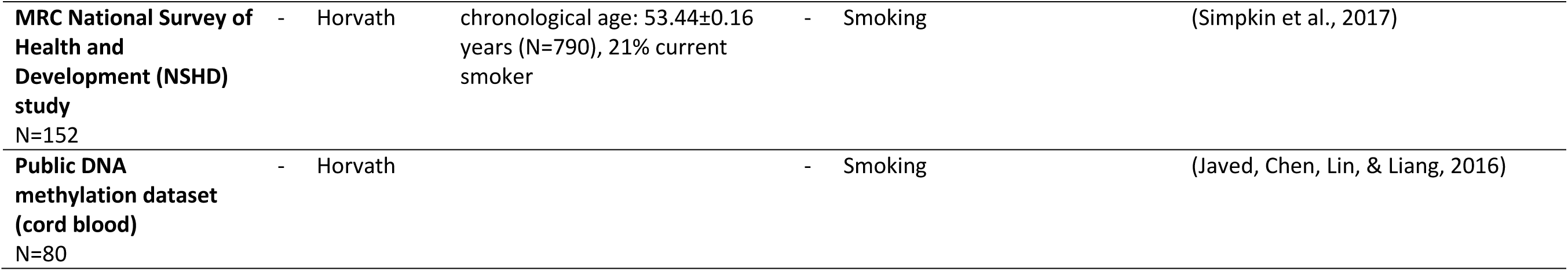
Overview of original work.

## Literature

Banszerus, V. L., Vetter, V. M., Salewsky, B., König, M., & Demuth, I. (2019). Exploring the Relationship of Relative Telomere Length and the Epigenetic Clock in the LipidCardio Cohort. International Journal of Molecular Sciences, 20(12), 1–14. https://doi.org/10.3390/ijms20123032

Beach, S. R. H., Dogan, M. V., Cutrona, C. E., Gerrard, M., Gibbons, F. X., Simons, R. L., … Philibert, R. A. (2015). Methylomic Aging As a Window on Lifestyle Impact: Tobacco and Alcohol Use Alter the Rate of Biological Aging Steven, 63(12), 2519–2525. https://doi.org/10.1002/jmri.25711.PET/MRI

Binder, A. M., Corvalan, C., Mericq, V., Pereira, A., Santos, L., Horvath, S., … Michels, K. B. (2018). Faster ticking rate of the epigenetic clock is associated with faster pubertal development in girls, 2294. https://doi.org/10.1080/15592294.2017.1414127

Busch, M. A., & Kuhnert, R. (2017). 12-Monats-Prävalenz einer koronaren Herzkrankheit in Deutschland. Journal of Health Monitoring, 2(1), 64–69. https://doi.org/10.17886/RKI-GBE-2017-009

Demuth, I., Bertram, L., Drewelies, J., Düzel, S., Lill, C. M., Lindenberger, U., … Gerstorf, D. (2019). Berlin Aging Study II (BASE-II). Encyclopedia of Gerontology and Population Aging, 1–8. https://doi.org/10.1007/978-3-319-69892-2_27-1

Dugué, P. A., Bassett, J. K., Joo, J. E., Baglietto, L., Jung, C. H., Wong, E. M., … Milne, R. L. (2018). Association of DNA Methylation-Based Biological Age with Health Risk Factors and Overall and Cause-Specific Mortality. American Journal of Epidemiology, 187(3), 529–538. https://doi.org/10.1093/aje/kwx291

Dugué, P., Bassett, J. K., Joo, J. E., Baglietto, L., Jung, C., Wong, E. M., … Milne, R. L. (2018). Original Contribution Association of DNA Methylation-Based Biological Age With Health Risk Factors and Overall and Cause-Specific Mortality, 187(3), 529–538. https://doi.org/10.1093/aje/kwx291

Ettehad, D., Emdin, C. A., Kiran, A., Anderson, S. G., Callender, T., Emberson, J., … Rahimi, K. (2016). Blood pressure lowering for prevention of cardiovascular disease and death: A systematic review and meta-analysis. The Lancet, 387(10022), 957–967. https://doi.org/10.1016/S0140-6736(15)01225-8

Gale, C. R., Marioni, R. E., Iva, Č., Chastin, S. F., Dall, P. M., Dontje, M. L., … Usp, S. (2018). The epigenetic clock and objectively measured sedentary and walking behavior in older adults?: the Lothian Birth Cohort 1936, 1–6. https://doi.org/10.1186/s13148-017-0438-z

Gao, X., Zhang, Y., Breitling, L. P., & Brenner, H. (2016). Relationship of tobacco smoking and smoking-related DNA methylation with epigenetic age acceleration, 7(30).

Hackshaw, A., Morris, J. K., Boniface, S., Tang, J. L., & Milenkovi, D. (2018). Low cigarette consumption and risk of coronary heart disease and stroke: Meta-analysis of 141 cohort studies in 55 study reports. BMJ (Online), 360. https://doi.org/10.1136/bmj.j5855

Hannum, G., Guinney, J., Zhao, L., Zhang, L., & Hughes, G. (2013). Genome-wide Methylation Profiles Reveal Quantitative Views of Human Aging Rates, 49(2), 359–367. https://doi.org/10.1016/j.molcel.2012.10.016.Genome-wide

Horvath, S. (2013). DNA methylation age of human tissues and cell types DNA methylation age of human tissues and cell types.

Horvath, S., Gurven, M., Levine, M. E., Trumble, B. C., Kaplan, H., Allayee, H., … Assimes, T. L. (2016). An epigenetic clock analysis of race/ethnicity, sex, and coronary heart disease. Genome Biology, 17(1), 0–22. https://doi.org/10.1186/s13059-016-1030-0

Horvath, S., Gurven, M., Levine, M. E., Trumble, B. C., Kaplan, H., Allayee, H., … Kobor, M. S. (2016). An epigenetic clock analysis of race / ethnicity, sex, and coronary heart disease An epigenetic clock analysis of race / ethnicity, sex, and coronary heart disease. Genome Biology, 0–22. https://doi.org/10.1186/s13059-016-1030-0

Irvin, M. R., Aslibekyan, S., Do, A., Zhi, D., Hidalgo, B., Claas, S. A., … Arnett, D. K. (2018). Metabolic and inflammatory biomarkers are associated with epigenetic aging acceleration estimates in the GOLDN study, 1–9.

Javed, R., Chen, W., Lin, F., & Liang, H. (2016). Infant’s DNA Methylation Age at Birth and Epigenetic Aging Accelerators, 2016.

Kaminsky, Z. A., Assadzadeh, A., Flanagan, J., & Petronis, A. (2005). Single nucleotide extension technology for quantitative site-specific evaluation of met C / C in GC-rich regions, 33(10), 1–12. https://doi.org/10.1093/nar/gni094

König, M., Joshi, S., Leistner, D. M., Landmesser, U., Sinning, D., Steinhagen-thiessen, E., & Demuth, I. (2019). Cohort profile?: role of lipoproteins in cardiovascular disease — the LipidCardio study. https://doi.org/10.1136/bmjopen-2019-030097

Kronenberg, F. (2016). Human Genetics and the Causal Role of Lipoprotein(a) for Various Diseases. Cardiovascular Drugs and Therapy, 30(1), 87–100. https://doi.org/10.1007/s10557-016-6648-3

Lee, J. S., Chang, P. Y., Zhang, Y., Kizer, J. R., Best, L. G., & Howard, B. V. (2017). Triglyceride and HDL-C dyslipidemia and risks of coronary heart disease and ischemic stroke by glycemic dysregulation status: The strong heart study. Diabetes Care, 40(4), 529–537. https://doi.org/10.2337/dc16-1958

Levine, M. E., Hosgood, H. D., Chen, B., Absher, D., Assimes, T., & Horvath, S. (2015). DNA methylation age of blood predicts future onset of lung cancer in the women’s health initiative, 7(9), 690–700.

Levine, M., Lu, A., Quach, A., Chen, B., Assimes, T., Bandinelli, S., … Horvath, S. (2018). An epigenetic biomarker of aging for lifespan and healthspan. BioRxiv.

Lind, L., Ingelsson, E., Sundstr, J., Siegbahn, A., & Lampa, E. (2017). Methylation-based estimated biological age and cardiovascular disease, (December), 1–7. https://doi.org/10.1111/eci.12872

Linke, A., Erbs, S., & Hambrecht, R. (2008). Effects of exercise training upon endothelial function in patients with cardiovascular disease. Frontiers in Bioscience, 13(6), 424–432.

Lu, J., Mu, Y., Su, Q., Shi, L., Liu, C., Zhao, J., … Ning, G. (2016). Reduced Kidney Function Is Associated With Cardiometabolic Risk Factors, Prevalent and Predicted Risk of Cardiovascular Disease in Chinese Adults: Results From the REACTION Study. Journal of the American Heart Association, 5(7). https://doi.org/10.1161/JAHA.116.003328

Luo, A., Jung, J., Longley, M., Rosoff, D. B., Charlet, K., Muench, C., … Lohoff, F. W. (2019). Epigenetic aging is accelerated in alcohol use disorder and regulated by genetic variation in APOL2. Neuropsychopharmacology, (August). https://doi.org/10.1038/s41386-019-0500-y

McCartney, D. L., Stevenson, A. J., Walker, R. M., Gibson, J., Morris, S. W., Campbell, A., … Marioni, R. E. (2018). Investigating the relationship between DNA methylation age acceleration and risk factors for Alzheimer’s disease. Alzheimer’s & Dementia: Diagnosis, Assessment & Disease Monitoring, 10, 429–437. https://doi.org/10.1016/j.dadm.2018.05.006

O’Neill, D., Britton, A., Hannah, M. K., Goldberg, M., Kuh, D., Khaw, K. T., & Bell, S. (2018). Association of longitudinal alcohol consumption trajectories with coronary heart disease: A meta-analysis of six cohort studies using individual participant data. BMC Medicine, 16(1), 1–13. https://doi.org/10.1186/s12916-018-1123-6

Pathak, L., Shirodkar, S., Ruparelia, R., & Rajebahadur, J. (2017). Coronary artery disease in women. Indian Heart Journal, (69), 532–538. https://doi.org/10.5114/kitp.2018.74675

Perna, L., Zhang, Y., Mons, U., Holleczek, B., Saum, K., & Brenner, H. (2016). Epigenetic age acceleration predicts cancer, cardiovascular, and all-cause mortality in a German case cohort. Clinical Epigenetics, 1–7. https://doi.org/10.1186/s13148-016-0228-z

Quach, A., Levine, M. E., Tanaka, T., Lu, A. T., Chen, B. H., Ferrucci, L., … Horvath, S. (2017). Epigenetic clock analysis of diet, exercise, education, and lifestyle factors. Aging, 9(2), 419–446. https://doi.org/10.18632/aging.101168

Quach, A., Levine, M. E., Tanaka, T., Lu, A. T., Chen, B. H., Ritz, B., … Eric, A. (2017). Epigenetic clock analysis of diet, exercise, education, and lifestyle factors, 9(2), 419–437.

Robert Koch Institut. (2015). Gesundheit in Deutschland (2015). Robert Koch-Institut (Hrsg) (2015) Gesundheit in Deutschland. Gesundheitsberichterstattung des Bundes. Gemeinsam getragen von RKI und Destatis. RKI, Berlin. https://doi.org/10.17886/rkipubl-2015-003-12

Roetker, N., Pankow, J., Bressler, J., Morrison, A., & Boerwinkler, E. (2018). Prospective Study of Epigenetic Age Acceleration and Incidence of Cardiovascular. Circ Genom Precis Med., 11 (March), 1–10. https://doi.org/10.1161/CIRCGEN.117.001937

Rosen, A. D., Robertson, K. D., Hlady, R. A., Muench, C., Horvath, S., Kaminsky, Z. A., … Philibert, R. (2018). DNA methylation age is accelerated in alcohol dependence. Translational Psychiatry. https://doi.org/10.1038/s41398-018-0233-4

Ryan, J., Wrigglesworth, J., Loong, J., Fransquet, P. D., & Woods, R. L. (2019). A Systematic Review and Meta-analysis of Environmental, Lifestyle, and Health Factors Associated With DNA Methylation Age. The Journals of Gerontology: Series A, XX(Xx), 1–14. https://doi.org/10.1093/gerona/glz099

Shrestha, D., Workalemahu, T., & Tekola-Ayele, F. (2019). Maternal dyslipidemia during early pregnancy and epigenetic ageing of the placenta. Epigenetics, 14(10), 1030–1039. https://doi.org/10.1080/15592294.2019.1629234

Sillanpää, E., Ollikainen, M., Kaprio, J., Wang, X., Leskinen, T., Kujala, U. M., & Törmäkangas, T. (2019). Leisure-time physical activity and DNA methylation age - A twin study. Clinical Epigenetics, 11(1), 1–8. https://doi.org/10.1186/s13148-019-0613-5

Simons, R., Lei, M. K., Beach, S. R. H., Philibert, R., Cutrona, C., Gibbons, F., & Barr, A. (2017). Economic Hardship and Biological Weathering: The Epigenetics of Aging in a U.S. Sample of Black Women. Physiology & Behavior, 176, 139–148. https://doi.org/10.1016/j.physbeh.2017.03.040

Simpkin, A. J., Cooper, R., Howe, L. D., Relton, C. L., Smith, G. D., Teschendorff, A., … Hardy, R. (2017). Are objective measures of physical capability related to accelerated epigenetic age? Findings from a British birth cohort. https://doi.org/10.1136/bmjopen-2017-016708

Teven, S., Affner, M. H., Eppo, S., Ehto, L., Apani, T., Önnemaa, R., … Aakso, L. (1998). MOR TALITY FROM CORONARY HEAR T DISEASE IN SUB JEC TS WITH AND WITHOUT TYPE 2 DIABETES MORTALITY FROM CORONARY HEART DISEASE IN SUBJECTS WITH TYPE 2 DIABETES AND IN NONDIABETIC SUBJECTS WITH AND WITHOUT PRIOR MYOCARDIAL INFARCTION A BSTRACT Background Typ. The New England Journal of Medicine, 339, 229–234. https://doi.org/10.1056/NEJM199807233390404

Vetter, V. M., Meyer, A., Karbasiyan, M., Steinhagen-thiessen, E., Hopfenmüller, W., & Demuth, I. (2018). Epigenetic Clock and Relative Telomere Length Represent Largely Different Aspects of Aging in the Berlin Aging Study II (BASE-II), XX(Xx), 1–6. https://doi.org/10.1093/gerona/gly184

Vidal-Bralo, L., Lopez-golan, Y., & Gonzalez, A. (2016). Simplified Assay for Epigenetic Age Estimation in Whole Blood of Adults, 7 (July), 1–7. https://doi.org/10.3389/fgene.2016.00126

Vidal-bralo, L., Lopez-golan, Y., Gonzalez, A., & Gonzalez, A. (2017). Corrigendum?: Simplified Assay for Epigenetic Age Estimation in Whole Blood of Adults, 8(April), 1–2. https://doi.org/10.3389/fgene.2017.00051

Wang, H., Naghavi, M., Allen, C., Barber, R. M., Carter, A., Casey, D. C., … Zuhlke, L. J. (2016). Global, regional, and national life expectancy, all-cause mortality, and cause-specific mortality for 249 causes of death, 1980–2015: a systematic analysis for the Global Burden of Disease Study 2015. The Lancet, 388(10053), 1459–1544. https://doi.org/10.1016/S0140-6736(16)31012-1

Weidner, C. I., Lin, Q., Koch, C. M., Eisele, L., Beier, F., Ziegler, P., … Wagner, W. (2014). Aging of blood can be tracked by DNA methylation changes at just three CpG sites Aging of blood can be tracked by DNA methylation changes at just three CpG sites.

Wilkins, E., Wilson, L., Wickramasinghe, K., & Bhatnagar, P. (2017). European Cardiovascular Disease Statistics 2017 edition. European Heart Network.

Zhao, W., Ammous, F., Ratli, S., Liu, J., Yu, M., Mosley, T. H., … Smith, J. A. (2019). Education and Lifestyle Factors Are Associated with DNA Methylation Clocks in Older African Americans.

